# A genome-first approach to rare variants in hypertrophic cardiomyopathy genes *MYBPC3* and *MYH7* in a medical biobank

**DOI:** 10.1101/2021.05.26.21257880

**Authors:** Joseph Park, Elizabeth A Packard, Michael G Levin, Renae L Judy, Regeneron Genetics Center, Scott M Damrauer, Sharlene M Day, Marylyn D Ritchie, Daniel J Rader

## Abstract

‘Genome-first’ approaches to analyzing rare variants can reveal new insights into human biology and disease. Because pathogenic variants are often rare, new discovery requires aggregating rare coding variants into ‘gene burdens’ for sufficient power. However, a major challenge is deciding which variants to include in gene burden tests. Pathogenic variants in *MYBPC3* and *MYH7* are well-known causes of hypertrophic cardiomyopathy (HCM), and focusing on these ‘positive control’ genes in a genome-first approach could help inform variant selection methods and gene burdening strategies for other genes and diseases. Integrating exome sequences with electronic health records among 41,759 participants in the Penn Medicine BioBank, we evaluated the performance of aggregating predicted loss-of-function (pLOF) and/or predicted deleterious missense (pDM) variants in *MYBPC3* and *MYH7* for gene burden phenome-wide association studies (PheWAS). The approach to grouping rare variants for these two genes produced very different results: pLOFs but not pDM variants in *MYBPC3* were strongly associated with HCM, whereas the opposite was true for *MYH7*. Detailed review of clinical charts revealed that only 38.5% of patients with HCM diagnoses carrying an HCM-associated variant in *MYBPC3* or *MYH7* had a clinical genetic test result. Additionally, 26.7% of *MYBPC3* pLOF carriers without HCM diagnoses had clear evidence of left atrial enlargement and/or septal/LV hypertrophy on echocardiography. Our study shows the importance of evaluating both pLOF and pDM variants for gene burden testing in future studies to uncover novel gene-disease relationships and identify new pathogenic loss-of-function variants across the human genome through genome-first analyses of healthcare-based populations.

## Introduction

‘Genome-first’ approaches, in which genetic variants of interest are first identified and then analyzed for association with phenotypes, can be used to inform the genetic basis of human disease and reveal new insights into gene function and human biology.^1^ Particularly when applied to medical biobanks consisting of healthcare populations with DNA sequencing data linked to extensive electronic health record (EHR) phenotype data, genome-first approaches allow for agnostic phenome-wide association studies (PheWAS) to determine the clinical impact of specific genetic variants.^2;3^ With the rising use of large-scale whole-exome sequencing (WES) and identification of many rare coding variants predicted to impact protein structure or function, studies are increasingly interrogating the cumulative effect of multiple rare variants in a gene (*i.e*. ‘gene burden’) to increase the statistical power of regression analyses and enable gene-based association studies to describe the implications of mutated genes in human disease.^4^

We have previously shown that gene-burden PheWAS applied to large healthcare populations has the potential to uncover novel consequences of rare coding variants in the human disease phenome.^5^ One approach to gene-burden PheWAS is to focus only on predicted loss-of-function (pLOF) variants, but this could lead to lack of power due to their infrequency. Additional coding variation could be added to substantially increase the number of effect alleles, but a major challenge is deciding which variants to include in gene burden tests of association. Furthermore, there are many *in silico* algorithms that can predict the probability that a variant may have a deleterious effect on its gene product as well as various filters that can be applied for variant selection based on variant type and frequency, but very little large-scale functional data that can be used to annotate missense variants.

Application of the unbiased genome-first approach to ‘positive control’ genes with known phenotype associations represents a valuable system for comparison of variant selection methods and gene burdening strategies, as we previously showed for the gene *LMNA* and its association with dilated cardiomyopathy.^6^ Pathogenic variants in *MYBPC3* and *MYH7* are known to cause hypertrophic cardiomyopathy (HCM) and together account for up to 50% of all clinically recognized HCM cases and at least 75% of HCM cases for which a pathogenic variant is identified.^7^ We leveraged the Penn Medicine BioBank (PMBB, University of Pennsylvania), a large academic medical biobank with WES linked to EHR data, to evaluate in detail the performance of methods for aggregating pLOF and/or annotated predicted deleterious missense (pDM) variants in *MYBPC3* and *MYH7* for gene burden association studies. Additionally, we followed up on our genome-first approach with review of EHR charts to describe the clinical characteristics of variant carriers identified through this gene burden study.

## Material and Methods

### Setting and study participants

All individuals recruited for the Penn Medicine BioBank (PMBB) are patients of clinical practice sites of the University of Pennsylvania Health System. Appropriate consent was obtained from each participant regarding storage of biological specimens, genetic sequencing, and access to all available EHR data. This study was approved by the Institutional Review Board of the University of Pennsylvania and complied with the principles set out in the Declaration of Helsinki.

### Whole-exome sequencing

This study included a subset of 43,731 individuals in the PMBB who had undergone whole-exome sequencing. We extracted DNA from stored buffy coats and then mapped exome sequences as generated by the Regeneron Genetics Center (Tarrytown, NY) to GRCh38 as previously described.^6^ Samples with low exome sequencing coverage, high missingness (*i.e*. greater than 5% of targeted bases), dissimilar reported and genetically determined sex, and genetic evidence of sample duplication were not included in this subset.^5;6^ For subsequent phenotypic association analyses, we removed samples with evidence of 1^st^ and 2^nd^-degree relatedness, leading to a total of sample size of 41,759 for analysis.

### Variant annotation and selection for gene-burden association testing

For PMBB, variants were annotated using ANNOVAR^8^ as pLOF or missense variants. pLOFs were defined as frameshift insertions or deletions, gain of stop codon, and disruption of canonical splice site dinucleotides. For splicing variants, we removed those with SpliceAI scores < 0.2 for loss or gain of acceptor or donor site.^9^ Several approaches to inclusion of rare variants in the gene burden were applied, including pLOFs only, additional ClinVar pathogenic variants, and inclusion of predicted deleterious missense (pDM) variants that were scored deleterious by 4/4 algorithms (SIFT^10^, PolyPhen2 HumDiv, Polyphen2 HumVar^11^, MutationTaster^12^). To capture additional individuals with potentially pathogenic missense variants, we utilized an ensemble method for predicting the pathogenicity of missense variants called REVEL^13^ to score rare missense variants in *MYBPC3* and *MYH7*. Finally, we overlapped a list of all 19 expert-adjudicated pathogenic missense variants in *MYBPC3* from the SHaRe Database to identify high-confidence pathogenic missense variants in PMBB.^14^

### Clinical data collection

All International Classification of Diseases Ninth Revision (ICD-9) and Tenth Revision (ICD-10) diagnosis codes, clinical imaging and laboratory measurements were extracted from the patients’ EHR. All ICD diagnosis codes and outpatient laboratory measurements available up to July 2020 were extracted for PMBB participants. Inpatient and outpatient echocardiography measurements were extracted if available for participants from September 2005 until November 2018. Outliers for each echocardiographic parameter (values >10 median absolute deviations from the median) were removed. Minimum, median, and maximum measurements of each quantitative trait were recorded per individual.

### Phenome-wide association studies

A PheWAS approach was used to determine the phenotypes associated with predicted deleterious variants in *MYBPC3* or *MYH7* carried by individuals in PMBB^15^. ICD-10 encounter diagnoses were mapped to ICD-9 via the Center for Medicare and Medicaid Services 2017 General Equivalency Mappings (https://www.cms.gov/Medicare/Coding/ICD10/2017-ICD-10-CM-and-GEMs.html) and manual curation. Phenotypes for each individual were then determined by mapping ICD-9 codes to distinct disease entities (*i.e*. Phecodes) using the R package “PheWAS”^16^. Patients were determined to be a case for a certain Phecode if they had the corresponding ICD diagnosis on 2 or more dates, while controls consisted of individuals who never had the ICD code. Individuals with an ICD diagnosis on only one date as well as individuals meeting control exclusion criteria based on default PheWAS phenotype mapping protocols were not considered in statistical analyses.

Each disease phenotype was tested for association with a gene burden of pLOF and/or pDM variants using a logistic regression model adjusted for age, sex, and the first ten principal components of genetic ancestry. We used an additive genetic model to aggregate variants into gene burdens as previously described.^5^ PheWAS analyses were performed separately by African and European genetic ancestry and then combined with inverse variance weighted meta-analysis. Our association analyses considered only disease phenotypes with at least 20 cases based on a prior simulation study for power analysis of rare variant gene burden PheWAS.^5^ This led to the interrogation of 1396 total Phecodes, and we used a Bonferroni correction to adjust for multiple testing (p=0.05/1396=3.58E-05).

### Statistical analyses

We also created a single HCM phenotype by combining Phecodes “Hypertrophic obstructive cardiomyopathy” and “Other hypertrophic cardiomyopathy” for association with various gene burdens in conjunction with PheWAS, using a logistic regression model adjusted for age, sex, and the first ten principal components of genetic ancestry. These HCM-specific analyses were performed separately by African and European genetic ancestry and combined with inverse variance weighted meta-analysis. Additionally, to compare available echocardiographic and serum laboratory measurements between carriers of predicted deleterious variants and genotypic controls, we used linear regression adjusted for age, sex, and the first ten principal components of genetic ancestry. These analyses were performed separately by African and European genetic ancestry and combined with inverse variance weighted meta-analysis. For echocardiographic comparison of *MYBPC3* pLOF and pathogenic missense variant carriers versus controls, *MYH7* pLOF and REVEL-informed pDM variant carriers were removed from controls. Likewise, for echocardiographic comparison of carriers of pLOF and pDM with REVEL≥0.5 variants in *MYH7* versus controls, carriers of *MYBPC3* pLOF and pathogenic missense variants were removed from controls. All statistical analyses were completed using R version 3.5 (Vienna, Austria).

### Review of clinical charts

We reviewed the clinical charts of patients with HCM who also carry a pLOF or pDM variant in *MYBPC3* or *MYH7* to assess the prevalence of clinical genetic testing for a molecular diagnosis of carrying a pathogenic *MYBPC3* or *MYH7* variant. We also reviewed the clinical charts of *MYH7* pLOF carriers to characterize a cardiac phenotype among these individuals without an HCM diagnosis. Finally, we interrogated the clinical charts of cases for “muscular wasting and disuse atrophy” who also carried a pLOF or pDM variant in *MYH7* to assess the prevalence of clinical genetic testing for a molecular diagnosis of *MYH7*-related myopathy.

## Results

### pLOF variants in MYBPC3 were strongly associated with HCM

Among 41,759 unrelated individuals with WES in PMBB (Table 1), we identified 45 individuals carrying one of 33 predicted loss-of-function (pLOF) variants in *MYBPC3*, including 13 frameshift insertions/deletions, 9 gain of stop codon, and 11 splicing variants disrupting canonical splice site dinucleotides (Figure 1A, Table S1). PheWAS of the gene burden of pLOF variants in *MYBPC3* showed phenome-wide significant associations with HCM and related cardiac phenotypes such as cardiac conduction disorders, heart failure, heart transplant/surgery, and use of cardiac defibrillator (Figure S1, Table 2). 15 of the 45 individuals with a rare pLOF had a clinical diagnosis of HCM (Phecodes “Hypertrophic obstructive cardiomyopathy” or “Other hypertrophic cardiomyopathy”) (Table S1). 12 of 33 pLOFs were annotated in ClinVar as pathogenic or likely pathogenic (P/LP), and of the 18 carriers of these P/LP variants, 6 had a clinical diagnosis of HCM. Of the 21 pLOF variants without a P/LP classification in ClinVar, 9 were carried by a total of 9 individuals with diagnoses of HCM (Table S1).

**Table 1:** Penn Medicine BioBank whole exome-sequenced cohort characteristics Basic demographic characteristics and representative Phecodes identified by gene burden PheWAS for *MYBPC3* and *MYH7*. Each characteristic is labeled with count data and percent prevalence where appropriate. Individuals were determined to be a case for a Phecode if they had the corresponding ICD diagnosis on two or more dates, while controls consisted of individuals who never had the ICD code. Individuals with an ICD diagnosis on only one date as well as those under control exclusion criteria based on Phecode mapping protocols were not considered. AFR-African, AMR-Mixed American, EAS-East Asian, EUR-European, SAS-South Asian

|  |  |
| --- | --- |
| **Basic demographics** |  |
| Total population, N | 41759 |
| Female, N (%) | 20731 (49.6) |
| Median age, years | 63 |
| **Genetically informed ancestry** |  |
| AFR, N (%) | 10217 (24.5) |
| AMR, N (%) | 572 (1.4) |
| EAS, N (%) | 672 (1.6) |
| EUR, N (%) | 29362 (70.3) |
| SAS, N (%) | 564 (1.4) |
| **Phecodes** |  |
| Primary/intrinsic cardiomyopathies, N (%) | 2912 (7.6) |
| Hypertrophic obstructive cardiomyopathy, N (%) | 199 (0.6) |
| Other hypertrophic cardiomyopathy, N (%) | 184 (0.5) |
| Cardiac dysrhythmias, N (%) | 12130 (35.5) |
| Atrial fibrillation, N (%) | 5885 (21.0) |
| Atrial flutter, N (%) | 2324 (9.5) |
| Paroxysmal ventricular tachycardia, N (%) | 1960 (8.2) |
| Ventricular fibrillation and flutter, N (%) | 467 (2.1) |
| Cardiac conduction disorders, N (%) | 5688 (20.5) |
| Cardiac pacemaker/device in situ, N (%) | 3177 (12.6) |
| Cardiac defibrillator in situ, N (%) | 1945 (8.1) |
| Congestive heart failure; nonhypertensive, N (%) | 6224 (16.9) |
| Heart transplant/surgery, N (%) | 792 (2.5) |
| Cardiac shunt/heart septal defect, N (%) | 547 (1.4) |
| Cardiogenic shock, N (%) | 188 (0.5) |
| Muscular wasting and disuse atrophy, N (%) | 52 (0.1) |

**Table 2:** Association of *MYBPC3* and *MYH7* gene burdens with HCM in PMBB Summary statistics for gene burden associations with HCM (combining Phecodes “Hypertrophic obstructive cardiomyopathy” and “Other hypertrophic cardiomyopathy”) in PMBB using ClinVar P/LP, pLOF only, missense only (pDM and/or adjudicated), and pLOF + missense variants in *MYBPC3* (top) and *MYH7* (bottom). Each gene burden association is reported as beta, standard error (SE), odds-ratio (OR), p value, the number of carriers for variants included in the gene burden, and the number of carriers having the HCM phenotype.

| ***MYBPC3*** |  |  |  |  |  |  |
| --- | --- | --- | --- | --- | --- | --- |
| **Gene Burden** | **Beta** | **SE** | **OR** | **P** | **Carrier N** | **HCM N** |
| ClinVar P/LP (pLOF + missense) | 4.177 | 0.453 | 65.161 | 3.15E-20 | 45 | 8 |
| pLOF only | 5.112 | 0.411 | 166.008 | 1.52E-35 | 45 | 15 |
| pDM (REVEL≥0.6) only | 0.908 | 0.316 | 2.48 | 4.05E-03 | 628 | 10 |
| Adjudicated missense (SHaRe) | 2.785 | 0.76 | 16.204 | 2.46E-04 | 22 | 2 |
| pLOF + pDM | 1.949 | 0.214 | 7.023 | 7.99E-20 | 673 | 25 |
| pLOF + adjudicated missense | 4.402 | 0.321 | 81.638 | 1.03E-42 | 67 | 17 |

**Table 2:** Association of *MYBPC3* and *MYH7* gene burdens with HCM in PMBB
| ***MYH7*** |  |  |  |  |  |  |
| --- | --- | --- | --- | --- | --- | --- |
| **Gene Burden** | **Beta** | **SE** | **OR** | **P** | **Carrier N** | **HCM N** |
| ClinVar P/LP (pLOF + missense) | 4.737 | 0.301 | 11.404 | 1.29E-55 | 77 | 22 |
| pLOF only | -10.658 | 315.401 | 2.35E-05 | 9.73E-01 | 27 | 0 |
| pDM (REVEL≥0.5) only | 2.059 | 0.183 | 7.839 | 2.01E-29 | 858 | 37 |
| pLOF + pDM | 2.033 | 0.183 | 7.635 | 9.23E-29 | 885 | 37 |

**Figure 1:**
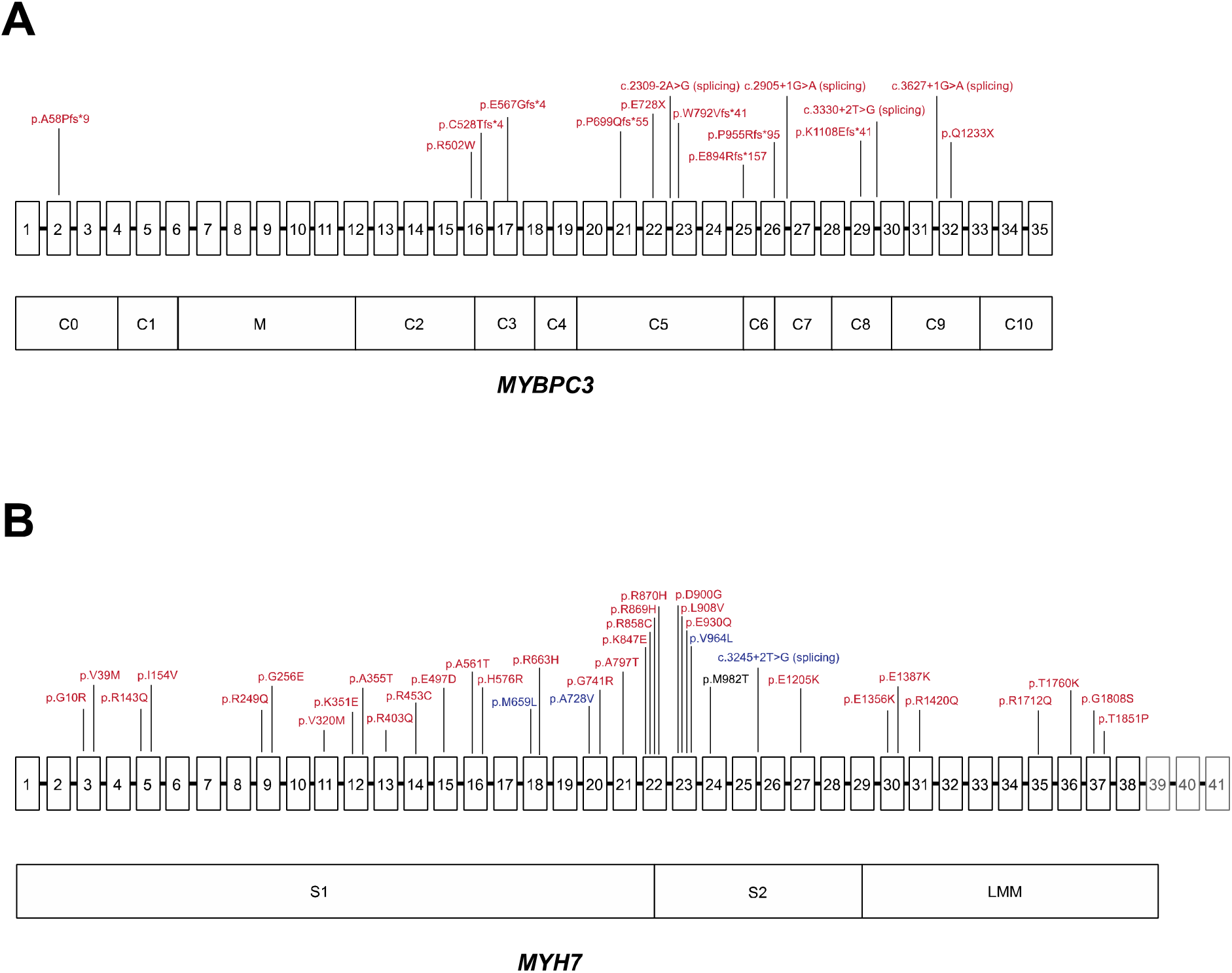
Distribution of disease-associated variants in *MYH7 and MYBPC3* A) Schematic of *MYBPC3* gene with exons 1-35 (exons not to scale) above and domains below, with variants labeled in red denoting amino acid change (or location of splice variant) for pLOF or adjudicated missense variants that were associated with HCM in PMBB. B) Schematic of *MYH7* gene with exons 1-41 (exons not to scale) above and domains below. Variants are labeled in red denoting amino acid change for pDM variants with REVEL≥0.5 that were associated with HCM in PMBB, and variants are labeled in blue denoting amino acid change (or location of splice variant) for pLOF or pDM variants with REVEL≥0.5 that were associated with “Muscular wasting and disuse atrophy” in PMBB. Note: exons 39-41, which do not encode for protein, are grayed out. Note 2: p.M982T in *MYH7* was associated with both HCM and muscular wasting and disuse atrophy, but via different individuals, and is thus labeled in black.

Review of clinical charts of the 15 pLOF carriers with a clinical diagnosis of HCM revealed that only 5 had a clinical genetic test report, all of which were concordant with the WES pLOF results. While all 5 individuals with clinical genetic testing had a family history of HCM, only 3 of 10 untested individuals had a family history of HCM noted in their charts. Chart review of the 30 *MYBPC3* pLOF carriers without a clinical HCM diagnosis revealed that 17 had received at least one transthoracic echocardiogram, and 8 of these individuals had left atrial enlargement and/or hypertrophy of the septum or another segment within the left ventricle. Additionally, 10 of 30 pLOF carriers without a diagnosis of HCM had a history of atrial fibrillation.

### Predicted deleterious missense (pDM) variants in MYBPC3 were not associated with HCM

Based on ‘phenotype-first’ presentations of HCM, most pathogenic variants in *MYBPC3* are pLOF variants.^17^ ClinVar shows that 482 frameshift, nonsense, or splicing variants in *MYBPC3* are classified as P/LP, whereas only 54 missense variants are annotated as P/LP.^18^ In PMBB, the gene burden of all nonsynonymous coding variants in *MYBPC3* classified by ClinVar as P/LP (N=45 heterozygous carriers) was strongly associated with HCM as expected (Figure S2, Table 2). However, while a gene burden of just the 12 pLOF variants classified as P/LP (N=18 heterozygous carriers) also showed phenome-wide significant associations with HCM Phecodes (Figure S3A), a gene burden of the 17 missense variants classified as P/LP (N=27 heterozygous carriers) showed a much weaker association with HCM that was not phenome-wide significant (p=9.50E-05) (Figure S3B). We previously have shown that prediction of deleteriousness for missense variants using the ensemble tool REVEL^13^ is highly correlated with clinical annotations for missense variants in *LMNA*.^6^ We similarly applied REVEL to missense variants in *MYBPC3* and found through an analysis of variance on ClinVar-annotated variants that REVEL scores showed essentially no correlation with annotations of clinical pathogenicity for *MYBPC3* (Table S2). We experimented with REVEL score thresholds in bins of 0.05 to evaluate the optimal score cutoff for capturing the most robust association of missense variants in *MYBPC3* associated with HCM. We found that all REVEL cutoff thresholds showed very weak association with HCM, although a REVEL threshold score of 0.6 was relatively optimal (Figure S4, Table 2). Additionally, application of the aggregation of predicted deleterious missense (pDM) variants chosen by a consensus of algorithms (SIFT, PolyPhen2 HumDiv, PolyPhen2 HumVar, and MutationTaster)—one of the standard approaches for predicting the deleteriousness of missense variants—also showed weak association with HCM (Figure S4).

Given the weak or insignificant association of missense variants in *MYBPC3* with HCM based on algorithms predicting deleteriousness of missense variants alone, we overlapped a list of 19 expert-adjudicated pathogenic missense variants in *MYBPC3* from the SHaRe Database to identify high-confidence pathogenic missense variants in PMBB.^14^ We identified 6 of these high-confidence pathogenic missense variants in *MYBPC3* carried by a total of 22 individuals among the PMBB WES cohort (Table S3). A gene burden including just these 6 missense variants showed a similar degree of association with HCM compared to a gene burden of ClinVar P/LP missense variants, and was more strongly associated with HCM compared to a burden of pDM variants based on all REVEL score thresholds as well as 4/4 algorithms (Figure S4, Table 2). 2 of the 22 carriers were diagnosed with HCM, and chart review of the 20 carriers without an HCM diagnosis revealed 5 individuals with mild concentric hypertrophy or dilated atria noted on transthoracic echocardiography.

Including pDMs together with pLOFs in a gene burden has the potential to increase power for finding phenotype associations. When we combined pLOFs with pDMs having REVEL≥0.6, we noted no improvement in the association of the gene burden with HCM (Table 2). Only when the adjudicated pathogenic missense variants were included with pLOFs did we see a stronger association with HCM compared with pLOFs alone (Figure 2, Table 2). We further interrogated this *MYBPC3* gene burden combining pLOFs and adjudicated pathogenic missense variants by analyzing available echocardiography data in PMBB. Carriers of pLOF and adjudicated pathogenic missense variants in *MYBPC3* on average had increased left ventricular posterior wall (LVPW) diastolic thickness, interventricular septum (IVS) diastolic thickness, left atrial (LA) volume index, and left ventricular outflow tract (LVOT) peak gradient (Table 3) compared to non-carriers, although not always at clinically relevant thresholds.

**Figure 2:**
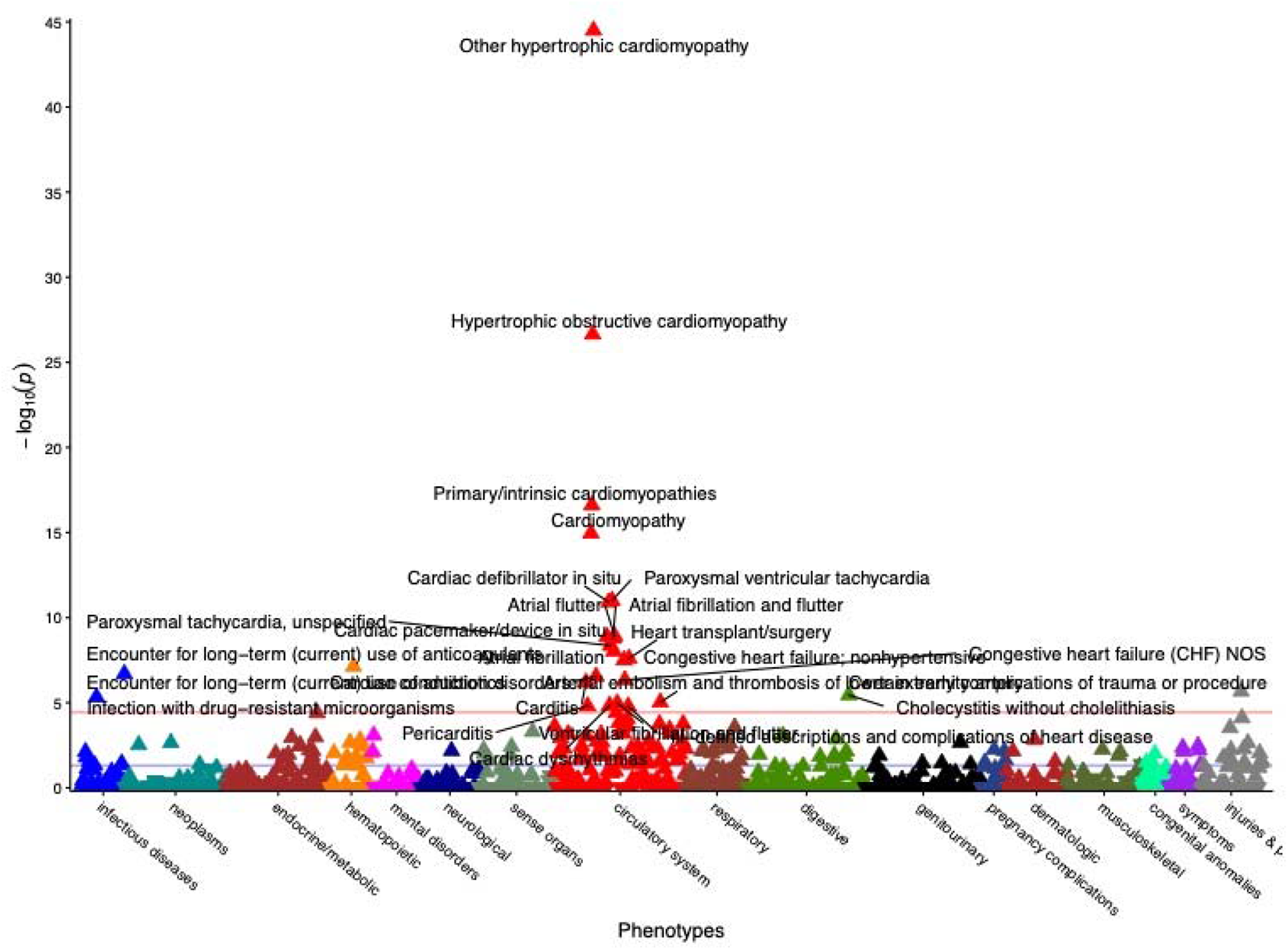
Gene burden PheWAS of pLOF and adjudicated pathogenic missense variants in *MYBPC3* Gene burden PheWAS of pLOF variants (N=45, Table S1) and adjudicated pathogenic missense variants from SHaRE (N=22, Table S3) in *MYBPC3*. Phecodes are plotted along the x axis to represent the phenome, and the association of the gene burden with each Phecode is plotted along the y axis representing –log_10_ (p value). The red line represents the Bonferroni-corrected significance threshold to adjust for multiple testing (p=3.58E-05), and the blue line represents a nominal significance threshold (p=0.05).

**Table 3:**
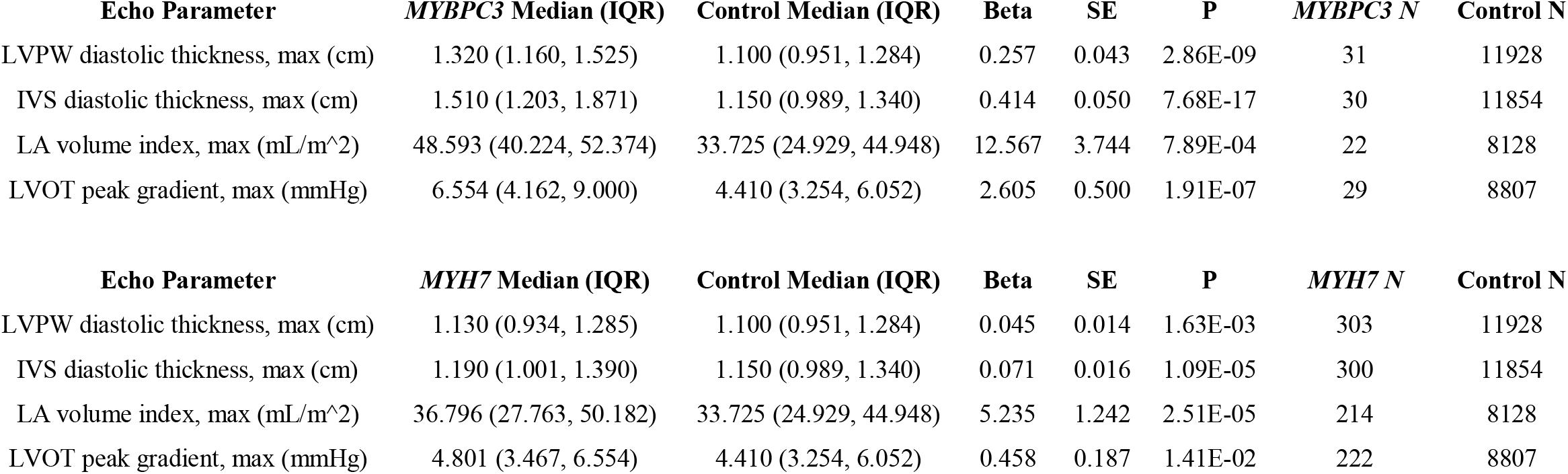
Echocardiographic analyses for *MYBPC3* and *MYH7* in PMBB <u>Top</u>: comparison of echocardiographic parameters representative of HCM with gene burden of pLOF variants and adjudicated pathogenic missense variants from SHaRE in *MYBPC3* versus non-carriers. Carriers of pLOF variants and pDM variants with REVEL≥0.5 in *MYH7* were removed from this analysis. Median and interquartile ranges (IQR) for each echo parameter are listed for each test group, and association results are listed as beta, standard error (SE), and p value. The number of individuals with each echo parameter available are also listed per test group. <u>Bottom</u>: comparison of echocardiographic parameters representative of HCM with gene burden of pLOF variants and pDM variants with REVEL≥0.5 in *MYH7* versus non-carriers. Carriers of pLOF variants and adjudicated pathogenic missense variants from SHaRE in *MYBPC3* were removed from this analysis. Median and interquartile ranges (IQR) for each echo parameter are listed for each test group, and association results are listed as beta, standard error (SE), and p value. The number of individuals with each echo parameter available are also listed per test group. LVPW – left ventricule posterior wall, IVS – interventricular septum, LA – left atrial, LVOT – left ventricular outflow tract

### pLOF variants in MYH7 were not significantly associated with HCM but had suggestive associations with other cardiac phenotypes

We identified 27 individuals carrying one of 23 pLOF variants in *MYH7*, including 5 frameshift insertion/deletions, 13 gain of stop codon, and 5 splicing variants (Table S4). Of note, none of these pLOF variants were in the last exon of *MYH7*. In contrast to *MYBPC3*, PheWAS of the *MYH7* pLOF only gene burden showed no association with HCM (p=0.973) (Figure S5, Table 2). We confirmed through chart review that none of the 27 pLOF carriers had an HCM diagnosis (Table S4). Interestingly, however, this pLOF gene burden had weak evidence for association with “Cardiac shunt/heart septal defect” (p=1.12E-04, N=3), suggesting the possibility of a cardiac phenotype among heterozygous carriers for pLOF variants in *MYH7*. Detailed review of the clinical charts of the 27 *MYH7* pLOF carriers confirmed a history of patent foramen ovale or atrial septal defect in 3 pLOF carriers, and also revealed that 13 of 27 had received echocardiograms, of which 5 individuals had mild concentric left ventricular hypertrophy.

### Predicted deleterious missense (pDM) variants in MYH7 were strongly associated with HCM

Most known pathogenic variants in *MYH7* are missense^19^, as exemplified by the ClinVar database which has classified 272 missense variants in *MYH7* as P/LP, whereas there are only 20 frameshift, nonsense, or splicing variants annotated as P/LP.^18^ In PMBB, we found a total of 43 missense variants annotated by ClinVar as P/LP (N=75 heterozygous carriers) as well as just two pLOFs annotated as P/LP (N=2 carriers). As expected, a gene burden comprised only of these ClinVar P/LP nonsynonymous variants had a very strong association with HCM (Figure S6, Table 2). Of the 75 carriers with ClinVar P/LP missense variants, 22 had a diagnosis of HCM, while neither of the two carriers for pLOFs annotated in ClinVar as P/LP had an HCM diagnosis.

We then asked how a computational approach to selecting *MYH7* missense variants would perform and explored in detail the association of predicted deleterious missense variants in *MYH7* with HCM and other phenotypes. We applied REVEL to missense variants in *MYH7* and found through an analysis of variance on ClinVar-annotated variants that REVEL scores were highly correlated with annotations of clinical pathogenicity for *MYH7* (Table S5). We then experimented with REVEL score thresholds in bins of 0.05 to evaluate the optimal score cutoff for capturing the most robust association of missense variants in *MYH7* with HCM, with the goal of potentially capturing deleterious missense variants in *MYH7* that lack a pathogenic ClinVar classification. We found that a REVEL cutoff score of 0.5 had the optimal p value for association of *MYH7* pDM variants with HCM (Figure S7, Table 2). Of note, this REVEL-based association was more strongly associated with HCM compared to the aggregation of pDM variants predicted deleterious by a consensus of algorithms (SIFT, PolyPhen2 HumDiv, PolyPhen2 HumVar, and MutationTaster), even while identifying more variants (303 variants with REVEL≥0.5 vs. 166 variants passing consensus of algorithms) (Figure S7). The HCM-prevalent missense variants were concentrated among the globular S1 head and coiled-coil S2 domains of *MYH7* (Figure 1B, Table S6). Among 858 total carriers for pDM variants in *MYH7* with REVEL≥0.5, 37 heterozygous carriers for 33 different pDM variants had a diagnosis of HCM (Table S6). 15 *MYH7* pDM variants with REVEL≥0.5 lacking a classification of P/LP in ClinVar were carried by individuals with a diagnosis of HCM (Table S6). Chart review of these 37 carriers confirmed the diagnosis of HCM and indicated that only 15 of the 37 carriers had clinical genetic testing for HCM in their chart (all were concordant with the WES *MYH7* variant). While 13 of 15 individuals with a clinical genetic test in the chart noted a family history of HCM, only half of untested individuals had a documented family history.

### Gene burden of pLOF and pDM variants in MYH7 was also associated with a skeletal muscle phenotype

In order to increase power for finding associations, we aggregated the 23 pLOF variants in *MYH7* (N=27 carriers) with the 303 pDM variants with REVEL≥0.5 in *MYH7* (N=858 carriers) into a gene burden. PheWAS of this combined *MYH7* gene burden showed, as expected, phenome-wide significant associations with HCM (Figure 3, Table 2), but the strength of this association was not greater than with a pDM gene burden alone. This expanded gene burden was also significantly associated with related cardiac phenotypes “Heart transplant/surgery” and “Cardiac defibrillator in situ” (Figure 3). We also linked the expanded gene burden with available echocardiography data in PMBB, and found that carriers of pLOF and pDM variants in *MYH7* on average had increased left ventricular posterior wall (LVPW) diastolic thickness, interventricular septum (IVS) diastolic thickness, left atrial (LA) volume index, and left ventricular outflow tract (LVOT) peak gradient (Table 3) compared to non-carriers, although not always at clinically relevant thresholds.

**Figure 3:**
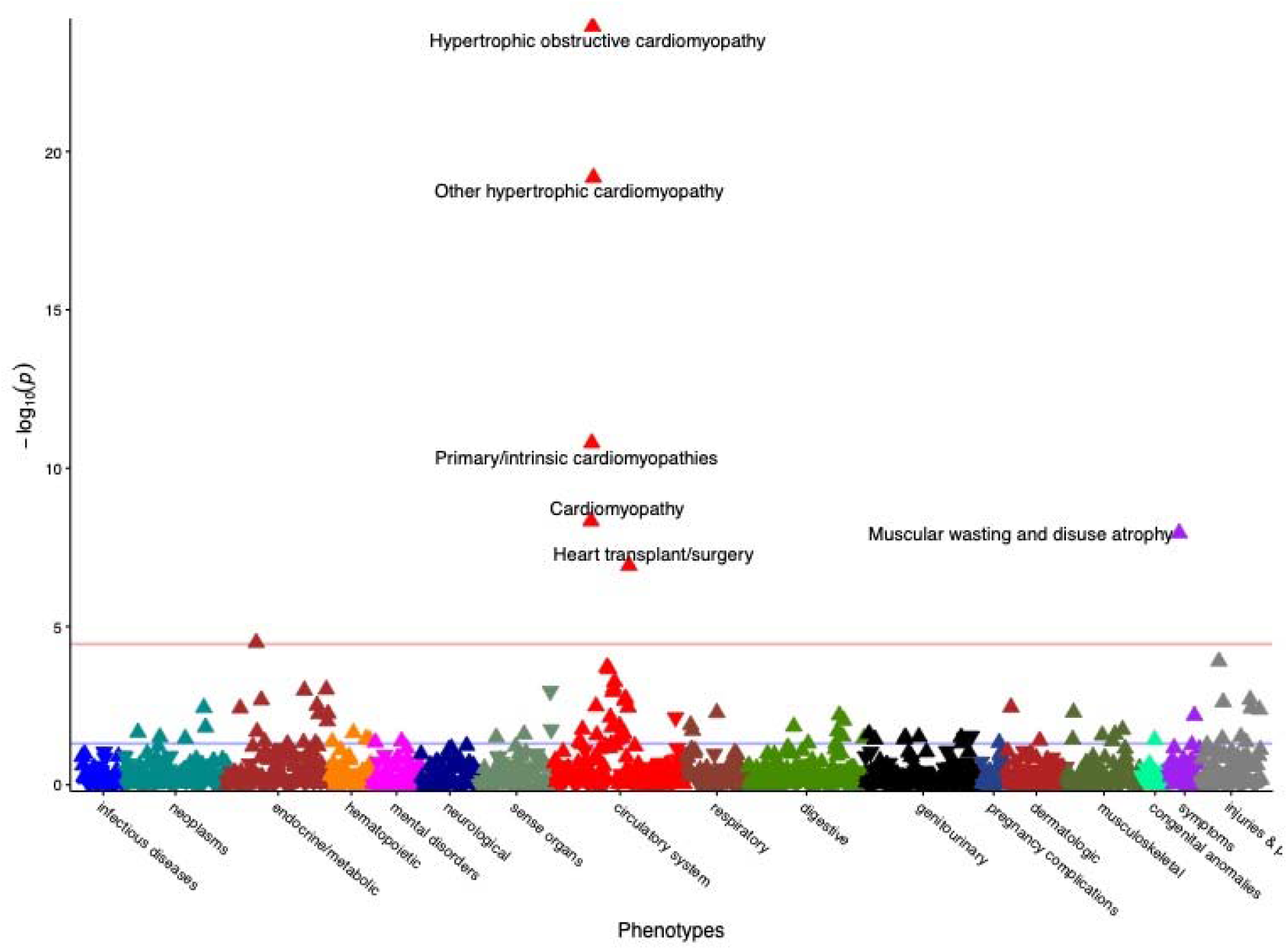
Gene burden PheWAS of pLOF + pDM variants in *MYH7* Gene burden PheWAS of pLOF variants (N=27, Table S4) and pDM variants with REVEL≥0.5 (N=858, Table S6) in *MYH7*. Phecodes are plotted along the x axis to represent the phenome, and the association of the gene burden with each Phecode is plotted along the y axis representing –log_10_ (p value). The red line represents the Bonferroni-corrected significance threshold to adjust for multiple testing (p=3.58E-05), and the blue line represents a nominal significance threshold (p=0.05).

Additionally, we found a phenome-wide significant association with “Muscular wasting and disuse atrophy” with this expanded gene burden (Figure 3) that was not seen with the gene burden limited to ClinVar P/LP variants alone (Figure S6). We identified 1 pLOF variant and 4 missense variants with REVEL≥0.5 carried by a total of 7 individuals who had the Phecode of “Muscular wasting and disuse atrophy” (Figure 1B, Table S4, Table S6). Of note, all 5 variants lacked a P/LP classification in ClinVar, and none of the 7 individuals had a diagnosis for HCM. Chart review of these individuals revealed secondary myopathy and generalized muscular wasting rather than a primary myopathy diagnosis. Of those individuals with electromyography performed, none revealed a primary neuromuscular diagnosis. We also compared available serum creatine kinase (CK) measurements between carriers for pLOF variants or pDM with REVEL≥0.5 in *MYH7* (total of 73 carriers with CK available) versus all non-carriers, and there was no significant association between the *MYH7* gene burden and CK concentrations (p=0.549).

## Discussion

Gene-burden PheWAS applied to large healthcare-based biobanks has the potential to elucidate the medical consequences of gene variants on the human disease phenome.^5^ A logical first step is to perform PheWAS focused only on predicted loss-of-function (pLOF) variants, which has the advantage of interrogating the largest effect sizes that a gene burden may have on associated phenotypes, but could lead to lack of power due to their infrequency. In aggregating pLOF variants in *MYBPC3* and *MYH7* for gene burden PheWAS in PMBB, only the *MYBPC3* pLOF gene burden was associated with HCM, while the *MYH7* pLOF gene burden was associated with other cardiac phenotypes and not HCM. This is an expected finding given that truncating variants account for >90% of cases of *MYBPC3* HCM,^20^ while only missense *MYH7* variants have a demonstrated association with HCM.^21^ Furthermore, detailed review of clinical charts confirmed a diagnosis of HCM among a subset of *MYBPC3* pLOF carriers while revealing mild concentric left ventricular hypertrophy and patent foramen ovale or atrial septal defect among a subset of *MYH7* pLOF carriers. Functional studies have shown that pLOF variants in *MYBPC3* promote nonsense-mediated decay pathways to contribute to the pathogenesis of HCM through haploinsufficiency.^17;22^ pLOF variants in *MYH7*, on the other hand, are not associated with HCM; however, recently pLOF variants in *MYH7* have been associated with left ventricular noncompaction cardiomyopathy, which has low diagnostic accuracy on echocardiography and thus could have been missed in our review of clinical charts.^23^

In principle, missense variants could be combined with pLOFs to substantially increase the number of effect alleles and power for novel discovery, but a major challenge is deciding which variants to include in gene burden tests of association. Including predicted deleterious missense (pDM) variants based on *in silico* prediction algorithms in addition to pLOFs in gene burdens increases power for disease associations in some genes, but the performance of such algorithms for missense variants may not be consistent across all genes. Our interrogation of missense variants based on prediction algorithms like REVEL serves as a testament to this notion; while pDM variants in *MYH7* showed strong correlation with clinical annotations of pathogenicity as well as strong associations with the expected HCM phenotype in PMBB, predictions of deleteriousness for missense variants in *MYBPC3* did not correlate with clinical annotations of pathogenicity and were not associated with HCM. The difference in performance may be due to different mechisms of HCM pathogenesis. For example, *MYH7* missense variants exert dominant negative effects on sarcomeric function, while haploinsufficiency and allelic imbalance in *MYBPC3* is the major mechanism leading to HCM.^24;25^

Importantly, these differences show that aggregating only pLOFs for gene burden association testing may be more appropriate for some genes like *MYBPC3*, while inclusion of additional pDM variants chosen based on *in silico* predictions may be more beneficial for increasing the number of effect alleles for others like *MYH7*, as we have similarly shown for *LMNA*.^6^ Additionally for *MYBPC3*, in which *in silico* prediction of missense variants performed poorly, we show that expert-adjudication of annotations of pathogenicity for missense variants based on *in vitro* and *in silico* analyses of missense variants in SHaRe, the largest comprehensive HCM cohort assembled to date, is superior in selecting missense variants for addition to gene burden testing.^14;20^ For other genes in which *in silico* predictions perform poorly for missense variants, we suggest that high-confidence pathogenic missense variants be added after expert adjudication to a degree that essentially parallels a saturated mutagenesis experiment.

A key goal in precision medicine initiatives is to promote a platform by which healthcare providers can make accurate diagnoses based on a wide variety of personalized health data, including individuals’ genetic information. Genetic testing of patients with HCM can identify the precise genetic cause of disease, improve diagnostic accuracy in a patient with an ambivalent diagnosis, and allow for cascade screening in family members.^26^ However, pathogenic HCM-causing variants found on more comprehensive sequencing can also be missed in clinical practice, which makes the genome-first approach applied to WES for review of clinical charts for carriers of pathogenic and potentially deleterious variants crucial.^27^ Our review of clinical charts revealed that a substantial number of patients clinically diagnosed with HCM did not appear to have had genetic testing, including those who were found in WES to have a pathogenic HCM-associated variant in *MYBPC3* or *MYH7*. We found that about a third of these HCM individuals without a genetic diagnosis were diagnosed with HCM before age 30. We also noted that patients with HCM without a genetic test in their chart had a significantly lower rate of a family history of HCM versus those who received genetic testing. While genetic testing is now routinely offered in specialized cardiomyopathy clinics, it is still broadly underutilized and patients may benefit from referral to specialized HCM clinics for genetic diagnosis and family screening.^28^ Additionally, we found that on average carriers for P/LP or predicted deleterious variants without a diagnosis of HCM had increased LA size and LV wall thickness compared to non-carriers, subclinical features that may warrant clinical follow-up when noted on echocardiography. Importantly, many carriers for P/LP or predicted deleterious variants without a diagnosis of HCM had no clinical or echocardiographic evidence of cardiac disease, showing the value of a genome-first approach for estimating penetrance of single-gene Mendelian disorders like HCM.

A major advantage of the genome-first approach to conducting gene burden PheWAS is the potential to capture pleiotropy. We found that the pLOF + pDM (REVEL ≥ 0.5) gene burden for *MYH7* had phenome-wide significant associations with both HCM and “Muscular wasting and disuse atrophy.” Importantly, while *MYBPC3* is specifically expressed in the heart, *MYH7* is also expressed in skeletal muscle,^29^ and *MYH7*-related myopathies are an emerging and underdiagnosed group of muscle diseases of childhood and adulthood.^30^ It has been reported that variants in *MYH7* which cause skeletal muscle disorders may cluster in the distal regions of the rod domain (light meromyosin domain, LMM with or without cardiac involvement).^30^ We found that myopathy-associated variants in *MYH7* were located in the neck and hinge (distal S1 and S2 domains) of *MYH7* from exons 18-24, suggesting that variants which affect skeletal muscle function may not be limited to the LMM domain. Of note, patients diagnosed with “muscular wasting and disuse atrophy” who carried a predicted deleterious *MYH7* variant did not have a primary myopathy diagnosis but rather had secondary muscular wasting, indicating that *MYH7-*related myopathy may be a ‘second hit’ phenomenon that can be unmasked by comorbidities.

In conclusion, we used a genome-first approach to include pLOFs and selected missense variants in *MYBPC3* and *MYH7* in gene burdens to show their significant associations with HCM, as well as the *MYH7*-specific association with myopathy. We demonstrate gene-specific differences in appropriate coding variant selection for gene burden testing to uncover important clinical and subclinical features relevant to associated diseases implicated by predicted deleterious variants. Our approach also suggests an expanded opportunity for clinical genetic evaluation and referral to multidisciplinary HCM centers. Importantly, our study demonstrates the value of assessing both pLOF and pDM variants for gene burden testing in future studies to uncover novel gene-disease relationships and identify novel pathogenic loss-of-function variants across the human genome through genome-first analyses of healthcare-based populations.

## Supporting information

Supplemental Figures (figures, titles, legends) and Tables (titles, legends)

## Data Availability

Up-to-date summary data for genetic variants captured via whole-exome sequencing in PMBB can be accessed via the Penn Medicine BioBank Genome Browser (https://pmbb.med.upenn.edu/biobank/allele-frequency/). Individual-level data are not made publicly available due to research participant privacy concerns; however, requests from accredited researchers for access to individual-level data relevant to this manuscript can be made by contacting the corresponding author.

## Supplemental Data

Supplemental Data include 7 figures and 6 tables.

## Acknowledgements

We thank JoEllen Weaver, Stephanie DerOhannessian, Marjorie Risman, Anurag Verma, Anastasia Lucas, and the Regeneron Genetics Center. The PMBB is funded by the Perelman School of Medicine at the University of Pennsylvania, a gift from the Smilow family, and the National Center for Advancing Translational Sciences of the National Institutes of Health under CTSA Award Number UL1TR001878. Research reported in this paper was supported by grants from the Blavatnik Family Foundation and National Human Genome Research Institute of the National Institutes of Health under award number F30HG010442 (to J.P.).

## References

1. Stessman, H.A., Bernier, R., and Eichler, E.E. (2014). A genotype-first approach to defining the subtypes of a complex disease. Cell 156, 872–877.

2. Dewey, F.E., Murray, M.F., Overton, J.D., Habegger, L., Leader, J.B., Fetterolf, S.N., O’Dushlaine, C., Van Hout, C.V., Staples, J., Gonzaga-Jauregui, C., et al. (2016). Distribution and clinical impact of functional variants in 50,726 whole-exome sequences from the DiscovEHR study. Science 354.

3. Bush, W.S., Oetjens, M.T., and Crawford, D.C. (2016). Unravelling the human genome-phenome relationship using phenome-wide association studies. Nat Rev Genet 17, 129–145.

4. Lee, S., Abecasis, G.R., Boehnke, M., and Lin, X. (2014). Rare-variant association analysis: study designs and statistical tests. Am J Hum Genet 95, 5–23.

5. Park, J., Lucas, A.M., Zhang, X., Chaudhary, K., Cho, J.H., Nadkarni, G., Dobbyn, A., Chittoor, G., Josyula, N.S., Katz, N., et al. (2021). Exome-wide evaluation of rare coding variants using electronic health records identifies new gene-phenotype associations. Nat Med 27, 66–72.

6. Park, J., Levin, M.G., Haggerty, C.M., Hartzel, D.N., Judy, R., Kember, R.L., Reza, N., Regeneron Genetics, C., Ritchie, M.D., Owens, A.T., et al. (2019). A genome-first approach to aggregating rare genetic variants in LMNA for association with electronic health record phenotypes. Genet Med.

7. Burke, M.A., Cook, S.A., Seidman, J.G., and Seidman, C.E. (2016). Clinical and Mechanistic Insights Into the Genetics of Cardiomyopathy. J Am Coll Cardiol 68, 2871–2886.

8. Wang, K., Li, M., and Hakonarson, H. (2010). ANNOVAR: functional annotation of genetic variants from high-throughput sequencing data. Nucleic Acids Res 38, e164.

9. Jaganathan, K., Kyriazopoulou Panagiotopoulou, S., McRae, J.F., Darbandi, S.F., Knowles, D., Li, Y.I., Kosmicki, J.A., Arbelaez, J., Cui, W., Schwartz, G.B., et al. (2019). Predicting Splicing from Primary Sequence with Deep Learning. Cell 176, 535–548 e524.

10. Kumar, P., Henikoff, S., and Ng, P.C. (2009). Predicting the effects of coding non-synonymous variants on protein function using the SIFT algorithm. Nat Protoc 4, 1073–1081.

11. Adzhubei, I.A., Schmidt, S., Peshkin, L., Ramensky, V.E., Gerasimova, A., Bork, P., Kondrashov, A.S., and Sunyaev, S.R. (2010). A method and server for predicting damaging missense mutations. Nat Methods 7, 248–249.

12. Schwarz, J.M., Cooper, D.N., Schuelke, M., and Seelow, D. (2014). MutationTaster2: mutation prediction for the deep-sequencing age. Nat Methods 11, 361–362.

13. Ioannidis, N.M., Rothstein, J.H., Pejaver, V., Middha, S., McDonnell, S.K., Baheti, S., Musolf, A., Li, Q., Holzinger, E., Karyadi, D., et al. (2016). REVEL: An Ensemble Method for Predicting the Pathogenicity of Rare Missense Variants. Am J Hum Genet 99, 877–885.

14. Thompson, A.D., Helms, A.S., Kannan, A., Yob, J., Lakdawala, N.K., Wittekind, S.G., Pereira, A.C., Jacoby, D.L., Colan, S.D., Ashley, E.A., et al. (2021). Computational prediction of protein subdomain stability in MYBPC3 enables clinical risk stratification in hypertrophic cardiomyopathy and enhances variant interpretation. Genet Med.

15. Denny, J.C., Bastarache, L., Ritchie, M.D., Carroll, R.J., Zink, R., Mosley, J.D., Field, J.R., Pulley, J.M., Ramirez, A.H., Bowton, E., et al. (2013). Systematic comparison of phenome-wide association study of electronic medical record data and genome-wide association study data. Nat Biotechnol 31, 1102–1110.

16. Carroll, R.J., Bastarache, L., and Denny, J.C. (2014). R PheWAS: data analysis and plotting tools for phenome-wide association studies in the R environment. Bioinformatics 30, 2375–2376.

17. Helms, A.S., Tang, V.T., O’Leary, T.S., Friedline, S., Wauchope, M., Arora, A., Wasserman, A.H., Smith, E.D., Lee, L.M., Wen, X.W., et al. (2020). Effects of MYBPC3 loss-of-function mutations preceding hypertrophic cardiomyopathy. JCI Insight 5.

18. Landrum, M.J., Lee, J.M., Benson, M., Brown, G.R., Chao, C., Chitipiralla, S., Gu, B., Hart, J., Hoffman, D., Jang, W., et al. (2018). ClinVar: improving access to variant interpretations and supporting evidence. Nucleic Acids Res 46, D1062–D1067.

19. Sabater-Molina, M., Perez-Sanchez, I., Hernandez Del Rincon, J.P., and Gimeno, J.R. (2018). Genetics of hypertrophic cardiomyopathy: A review of current state. Clinical genetics 93, 3–14.

20. Helms, A.S., Thompson, A.D., Glazier, A.A., Hafeez, N., Kabani, S., Rodriguez, J., Yob, J.M., Woolcock, H., Mazzarotto, F., Lakdawala, N.K., et al. (2020). Spatial and Functional Distribution of MYBPC3 Pathogenic Variants and Clinical Outcomes in Patients With Hypertrophic Cardiomyopathy. Circ Genom Precis Med 13, 396–405.

21. Walsh, R., Thomson, K.L., Ware, J.S., Funke, B.H., Woodley, J., McGuire, K.J., Mazzarotto, F., Blair, E., Seller, A., Taylor, J.C., et al. (2017). Reassessment of Mendelian gene pathogenicity using 7,855 cardiomyopathy cases and 60,706 reference samples. Genet Med 19, 192–203.

22. Seeger, T., Shrestha, R., Lam, C.K., Chen, C., McKeithan, W.L., Lau, E., Wnorowski, A., McMullen, G., Greenhaw, M., Lee, J., et al. (2019). A Premature Termination Codon Mutation in MYBPC3 Causes Hypertrophic Cardiomyopathy via Chronic Activation of Nonsense-Mediated Decay. Circulation 139, 799–811.

23. Klaassen, S., Probst, S., Oechslin, E., Gerull, B., Krings, G., Schuler, P., Greutmann, M., Hurlimann, D., Yegitbasi, M., Pons, L., et al. (2008). Mutations in sarcomere protein genes in left ventricular noncompaction. Circulation 117, 2893–2901.

24. Witjas-Paalberends, E.R., Piroddi, N., Stam, K., van Dijk, S.J., Oliviera, V.S., Ferrara, C., Scellini, B., Hazebroek, M., ten Cate, F.J., van Slegtenhorst, M., et al. (2013). Mutations in MYH7 reduce the force generating capacity of sarcomeres in human familial hypertrophic cardiomyopathy. Cardiovasc Res 99, 432–441.

25. Glazier, A.A., Thompson, A., and Day, S.M. (2019). Allelic imbalance and haploinsufficiency in MYBPC3-linked hypertrophic cardiomyopathy. Pflugers Arch 471, 781–793.

26. Cirino, A.L., Seidman, C.E., and Ho, C.Y. (2019). Genetic Testing and Counseling for Hypertrophic Cardiomyopathy. Cardiol Clin 37, 35–43.

27. Bagnall, R.D., Ingles, J., Dinger, M.E., Cowley, M.J., Ross, S.B., Minoche, A.E., Lal, S., Turner, C., Colley, A., Rajagopalan, S., et al. (2018). Whole Genome Sequencing Improves Outcomes of Genetic Testing in Patients With Hypertrophic Cardiomyopathy. J Am Coll Cardiol 72, 419–429.

28. Mital, S., Musunuru, K., Garg, V., Russell, M.W., Lanfear, D.E., Gupta, R.M., Hickey, K.T., Ackerman, M.J., Perez, M.V., Roden, D.M., et al. (2016). Enhancing Literacy in Cardiovascular Genetics: A Scientific Statement From the American Heart Association. Circ Cardiovasc Genet 9, 448–467.

29. Consortium, G.T. (2013). The Genotype-Tissue Expression (GTEx) project. Nat Genet 45, 580–585.

30. Fiorillo, C., Astrea, G., Savarese, M., Cassandrini, D., Brisca, G., Trucco, F., Pedemonte, M., Trovato, R., Ruggiero, L., Vercelli, L., et al. (2016). MYH7-related myopathies: clinical, histopathological and imaging findings in a cohort of Italian patients. Orphanet J Rare Dis 11, 91.

