## Supplemental Figures (figures, titles, legends) and Tables (titles, legends) for "A genome-first approach to rare variants in hypertrophic cardiomyopathy genes *MYBPC3* and *MYH7* in a medical biobank"

### Supplemental Figures and Tables

**Supplemental Figures**

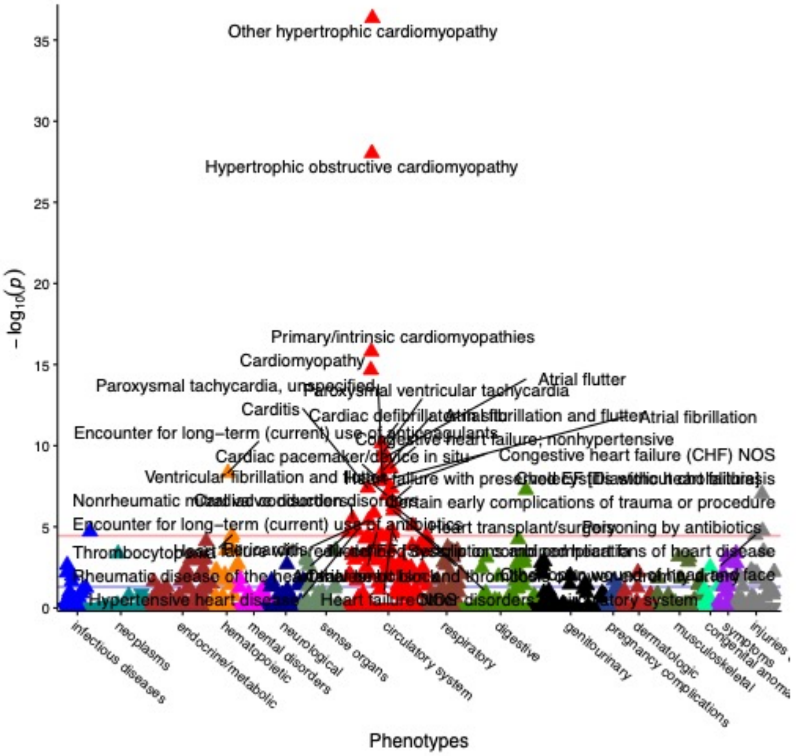


**Figure S1:** Gene burden PheWAS of pLOF variants in *MYBPC3*

Gene burden PheWAS of pLOF variants (N=45, Table S1) in *MYBPC3*. Phecodes are plotted along the x axis to represent the phenome, and the association of the gene burden with each Phecode is plotted along the y axis representing $-\log_{10}$(p value). The red line represents the Bonferroni-corrected significance threshold to adjust for multiple testing (p=3.58E-05), and the blue line represents a nominal significance threshold (p=0.05).

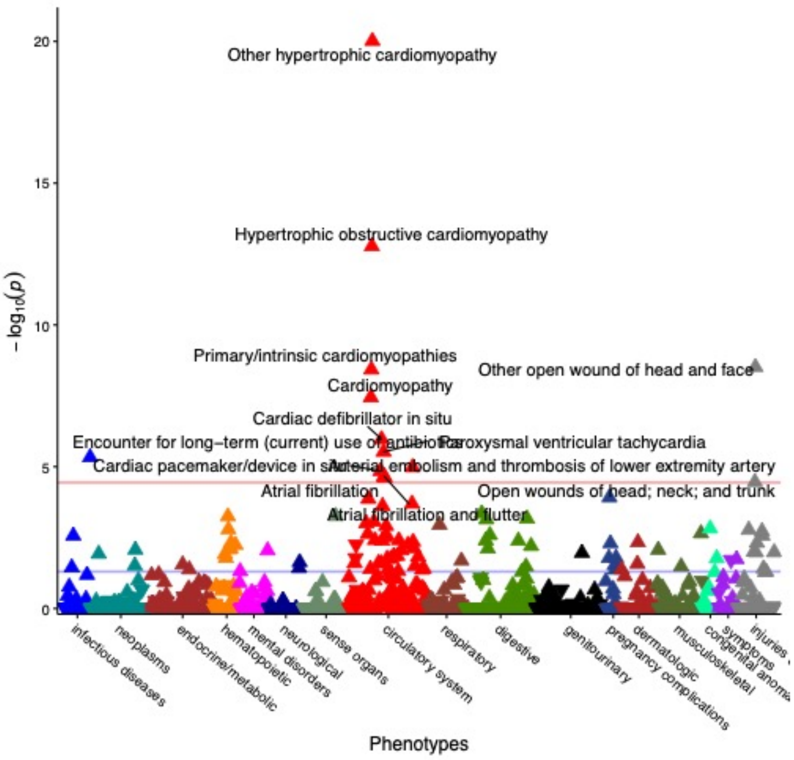


**Figure S2:** Gene burden PheWAS of ClinVar P/LP variants in *MYBPC3*

Gene burden PheWAS of nonsynonymous coding variants classified in ClinVar as P/LP in *MYBPC3* (N=45). Phecodes are plotted along the x axis to represent the phenome, and the association of the gene burden with each Phecode is plotted along the y axis representing $-\log_{10}$(p value). The red line represents the Bonferroni-corrected significance threshold to adjust for multiple testing (p=3.58E-05), and the blue line represents a nominal significance threshold (p=0.05).

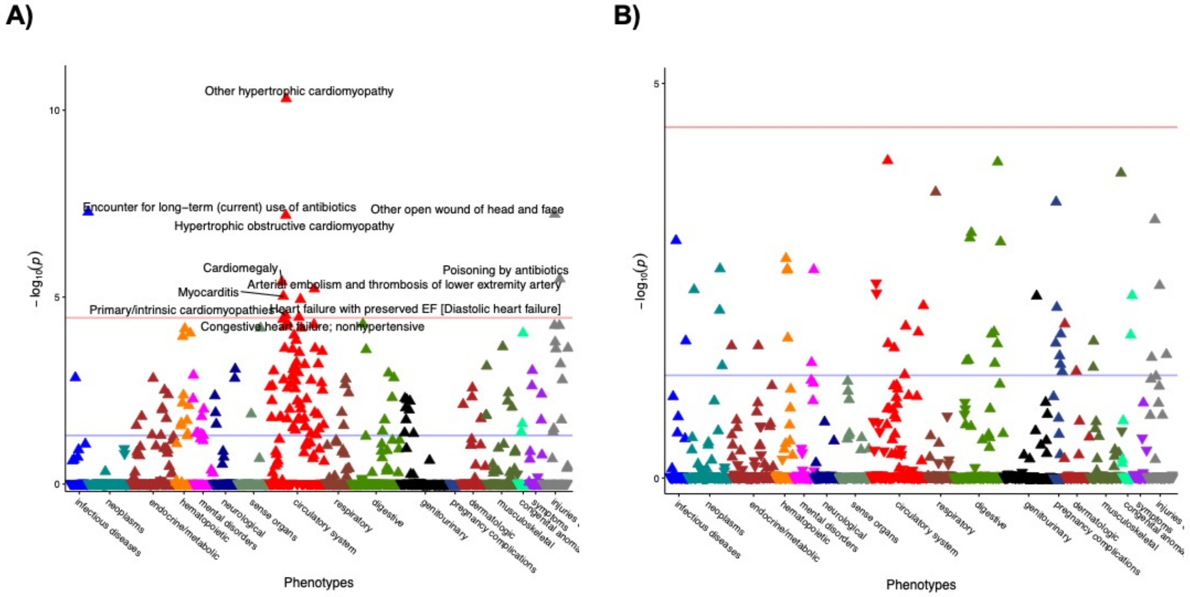


**Figure S3:** Gene burden PheWAS of ClinVar P/LP pLOF variants vs. ClinVar P/LP missense variants in *MYBPC3*

A) Gene burden PheWAS of pLOF variants in *MYBPC3* classified in ClinVar as P/LP (N=18), and B) gene burden PheWAS of missense variants in *MYBPC3* classified in ClinVar as P/LP (N=27). Phecodes are plotted along the x axis to represent the phenome, and the association of the gene burden with each Phecode is plotted along the y axis representing $-\log_{10}$(p value). The red line represents the Bonferroni-corrected significance threshold to adjust for multiple testing (p=3.58E-05), and the blue line represents a nominal significance threshold (p=0.05).

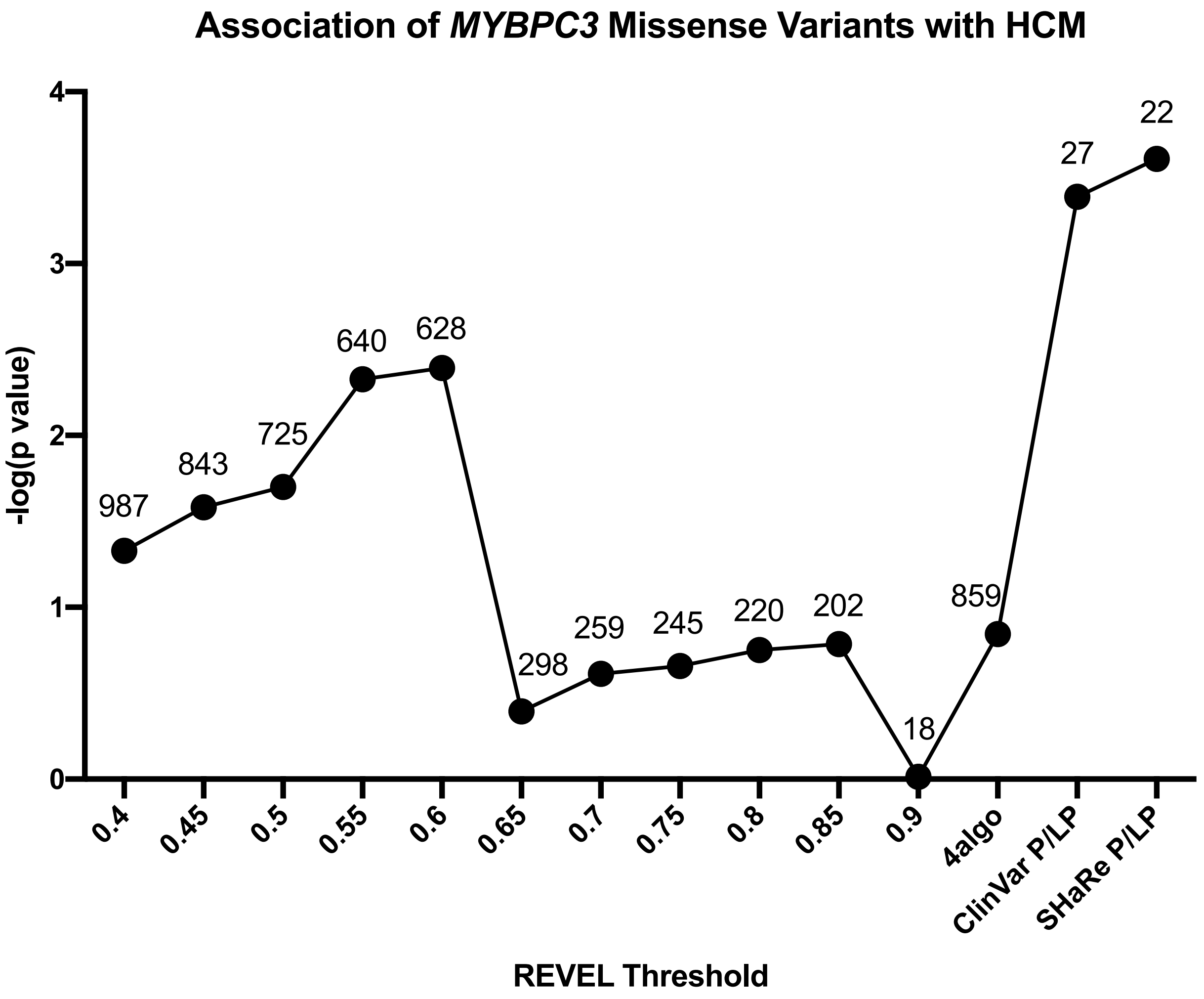


**Figure S4:** Selection of pDM variants in *MYBPC3* for association with HCM

Plot of p values for gene burden associations with HCM (combining Phecodes "Hypertrophic obstructive cardiomyopathy" and "Other hypertrophic cardiomyopathy") using missense variants in *MYBPC3* predicted to be deleterious per various REVEL cutoff scores as well as a consensus of algorithms (SIFT, PolyPhen2 HumDiv, PolyPhen2 HumVar, MutationTaster). Also included are missense variants classified as P/LP in ClinVar and SHaRe. Each point is labeled with the number of exome-sequenced individuals who are carriers for missense variants in each threshold category.

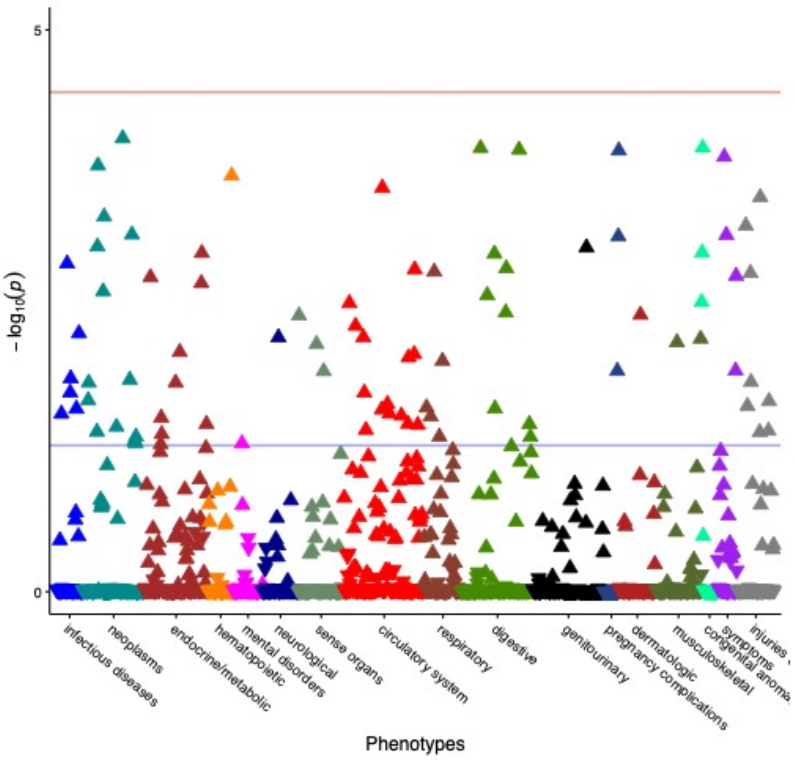


**Figure S5:** Gene burden PheWAS of pLOF variants *MYH7*

Gene burden PheWAS of pLOF variants (N=27, Table S4) in *MYH7*. Phecodes are plotted along the x axis to represent the phenome, and the association of the gene burden with each Phecode is plotted along the y axis representing $–\log_{10}$(p value). The red line represents the Bonferroni-corrected significance threshold to adjust for multiple testing (p=3.58E-05), and the blue line represents a nominal significance threshold (p=0.05).

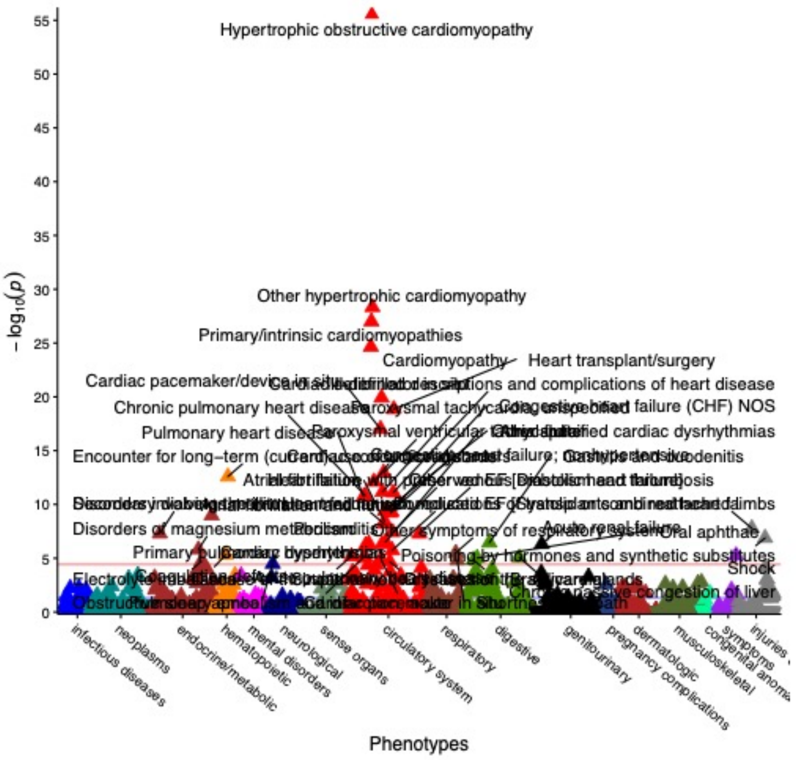


**Figure S6:** Gene burden PheWAS of ClinVar P/LP variants in *MYH7*

Gene burden PheWAS of nonsynonymous coding variants in *MYH7* classified in ClinVar as P/LP (N=77). Phecodes are plotted along the x axis to represent the phenome, and the association of the gene burden with each Phecode is plotted along the y axis representing $-\log_{10}$(p value). The red line represents the Bonferroni-corrected significance threshold to adjust for multiple testing (p=3.58E-05), and the blue line represents a nominal significance threshold (p=0.05).

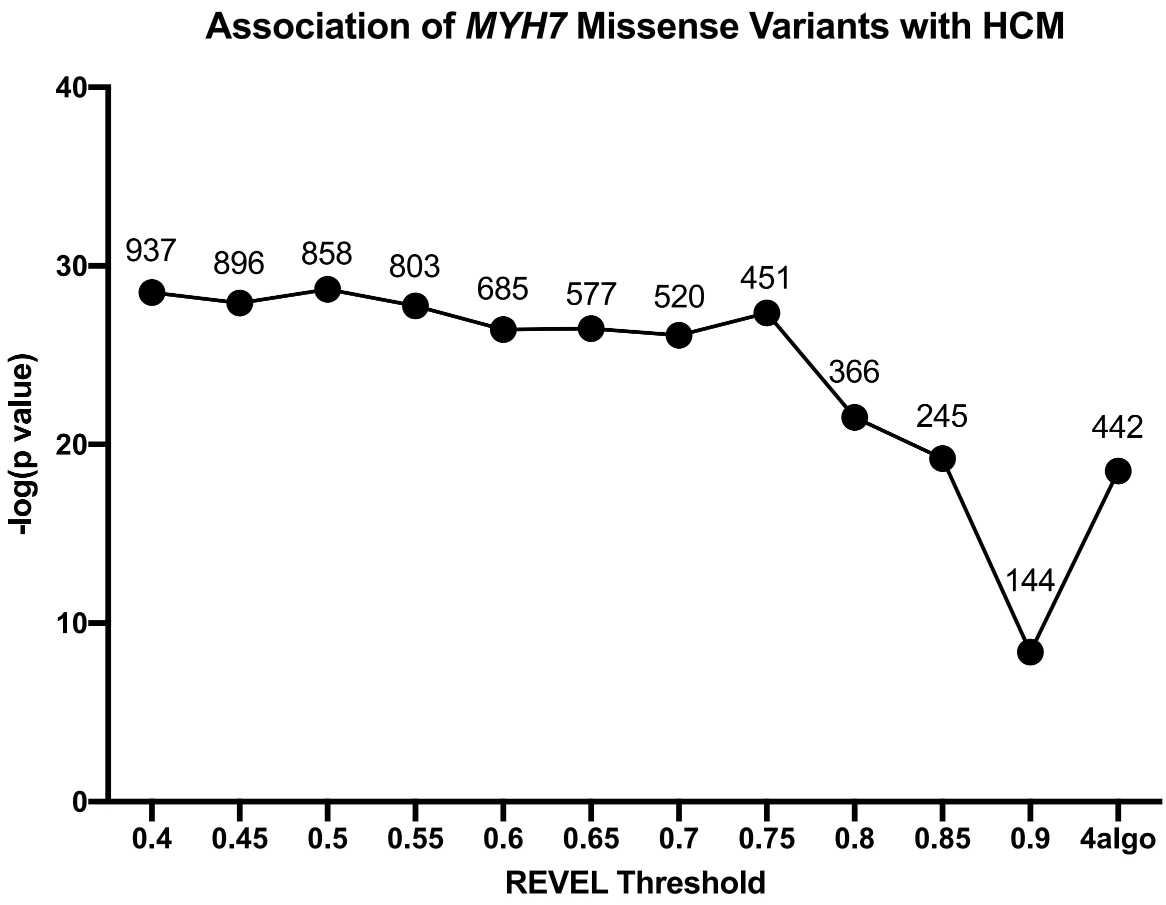


**Figure S7:** Selection of pDM variants in *MYH7* for association with HCM

Plot of p values for gene burden associations with HCM (combining Phecodes "Hypertrophic obstructive cardiomyopathy" and "Other hypertrophic cardiomyopathy") using missense variants in *MYH7* predicted to be deleterious per various REVEL cutoff scores as well as a consensus of algorithms (SIFT, PolyPhen2 HumDiv, PolyPhen2 HumVar, MutationTaster). Each point is labeled with the number of exome-sequenced individuals who are carriers for missense variants in each threshold category.

### Supplemental Table Titles and Legends

**Table S1:** List of pLOF variants in *MYBPC3* in PMBB

List of pLOF variants in *MYBPC3* found in the whole exome sequenced-cohort in PMBB. Each variant is annotated with chromosomal location per GRCh38, reference and alternate alleles, variant effect, exon number, amino acid (AA) change where relevant, ClinVar annotation if available, the number of carriers for the variant, and the number of carriers also diagnosed with HCM.

**Table S2:** Comparison of REVEL and ClinVar for missense variants in *MYBPC3*

Non-parametric one-way ANOVA on ranks (i.e. Kruskal-Wallis test by ranks) between ClinVar annotation categories and REVEL scores for missense variants in *MYBPC3*. P values from post-hoc analysis via the Dunn test are listed for all multiple comparisons of REVEL scores between P/LP and non-P/LP ClinVar categories.

**Table S3:** List of adjudicated pathogenic missense variants in *MYBPC3* in PMBB

List of missense variants in *MYBPC3* adjudicated as pathogenic or likely pathogenic in SHaRe found in the whole exome sequenced-cohort in PMBB. Each variant is annotated with chromosomal location per GRCh38, reference and alternate alleles, variant effect, exon number, amino acid (AA) change, ClinVar annotation if available, the number of carriers for the variant, and the number of carriers also diagnosed with HCM.

**Table S4:** List of pLOF variants in *MYH7* in PMBB

List of pLOF variants in *MYH7* found in the whole exome sequenced-cohort in PMBB. Each variant is annotated with chromosomal location per GRCh38, reference and alternate alleles, variant effect, exon number, amino acid (AA) change where relevant, ClinVar annotation if available, the number of carriers for the variant, the number of carriers also diagnosed with HCM, and the number of carriers also diagnosed with muscular wasting and disuse atrophy.

**Table S5:** Comparison of REVEL and ClinVar for missense variants in *MYH7*

Non-parametric one-way ANOVA on ranks (i.e. Kruskal-Wallis test by ranks) between ClinVar annotation categories and REVEL scores for missense variants in *MYH7*. P values from post-hoc analysis via the Dunn test are listed for all multiple comparisons of REVEL scores between P/LP and non-P/LP ClinVar categories.

**Table S6:** List of pDM variants with REVEL≥0.5 in *MYH7* in PMBB

List of pDM variants with REVEL≥0.5 in *MYH7* found in the whole exome sequenced-cohort in PMBB. Each variant is annotated with chromosomal location per GRCh38, reference and alternate alleles, variant effect, exon number, amino acid (AA) change, ClinVar annotation if available, the number of carriers for the variant, the number of carriers also diagnosed with HCM, and the number of carriers also diagnosed with muscular wasting and disuse atrophy.
